# Evaluation of the ratio of the transverse diameter of cerebellum to the transverse diameter of frontal lobe by trans-cerebellar section

**DOI:** 10.1101/2021.05.07.21256820

**Authors:** Xu Pingping, Zhang Dirong, Shi Yu, Kong Fengbei, Yao Chunxiao, She Ying, Wu Guoru

## Abstract

**Objective:** On the basis of a prospective study of evaluating the development of frontal lobe in mid-to-late trimester by trans-cerebellar section, it had been found that the transverse frontal diameter had a higher correlation with gestational age than the anteroposterior frontal diameter. In order to provide more valuable information for the prenatal diagnosis of suspected fetal microcephaly, the reference value range of the ratio of the transverse diameter of cerebellum to the transverse diameter of frontal lobe by trans-cerebellar section was further prospectively established.

**Methods:** A prospective cross-sectional study was conducted on 870 normal fetuses at 21 to 36 weeks of gestation from January 2019 to June 2020. The transverse diameter of cerebellum and the transverse diameter of frontal lobe were measured by trans-cerebellar section, and the ratio of the transverse diameter of cerebellum to the transverse diameter of frontal lobe was calculated, and the correlation was analyzed statistically.

**Results:** The ratio of the transverse diameter of cerebellum to the transverse diameter of frontal lobe increased positively with gestational age which correlation coefficient was 0.413 (P < 0.001). Taking gestational age as the independent variable and the ratio of the transverse diameter of cerebellum to the transverse diameter of frontal lobe as the dependent variable, it showed that there was a linear relationship between the gestational age and the ratio of the transverse diameter of cerebellum to the transverse diameter of frontal lobe. We got a correlation formula that the ratio of the transverse diameter of cerebellum to the transverse diameter of frontal lobe =0.004× (gestational age-21)+0.530.

**Conclusion:** This study has preliminarily established the normal reference range of the ratio of the transverse diameter of cerebellum to the transverse diameter of frontal lobe by trans-cerebellar section in normal mid-to-late pregnancy fetuses, which can provide valuable information for the prenatal diagnosis of suspected fetal microcephaly.

## INTRODUCTION

Microcephaly is a developmental disorder of the nervous system. Pathologically, microcephaly is mainly manifested by developmental retardation of the frontal lobe. Clinically, microcephaly is mainly manifested by small head circumference, especially the reduction of the forehead. Severe cases may be manifested as special facial features such as a backward tilt of the forehead and mental retardation^1^. At present, there is no gold standard for prenatal diagnosis of microcephaly. That fetal head circumference is less than three standard deviations of the mean value of normal fetuses in the same gestational week is the clue for prenatal ultrasound diagnosis of microcephaly^2^. However, the diagnosis of fetal microcephaly only based on this index has a high false positive rate which is up to 43%^3^. Since the frontal lobe of the brain is more involved than the cerebellum in microcephaly, the ratio of cerebellum to frontal lobe related measurement parameters in microcephaly will increase theoretically. However, the relationship is not clear between the ratio of cerebellum to frontal lobe related development parameters and gestational age in fetal period. In our previous study, we have found that the transverse diameter of fetal frontal lobe is more correlated with gestational age than the anteroposterior frontal diameter. On this basis, this study aims to further establish the normal reference range of the ratio of the transverse diameter of cerebellum to the transverse diameter of frontal lobe by trans-cerebellar section in normal mid-to-late pregnancy fetuses so as to provide valuable information for the prenatal diagnosis of fetal microcephaly.

## METHODS

This was a prospective cross-sectional study. Normal fetuses undergoing routine prenatal ultrasonography in our hospital from January 2019 to June 2020 were selected as subjects. To exclude abnormal fetuses and ensure the relative accuracy of gestational age, the inclusion criteria were as follows: the date of the last menstruation was clear or the gestational week was corrected by crown-lump length;21 to 36 weeks of gestation; low risk pregnancy including pregnant women without diabetes, hypertension and other pregnancy risk factors. Exclusion criteria were as follows: fetuses with known malformations or chromosomal abnormalities; the large difference between the biological gestational age and the actual gestational age of the fetuses; multiple pregnancies; nervous system abnormality occurs within 6 months after delivery.

This study was approved by the Peking University Shenzhen Hospital and Health Service Human Research Ethics Committee. All pregnant women gave informed consent to this study. The pregnant women ranged in age from 20 to 39 years old, with an average age of 27 years old. The gestational age ranged from 21 to 36weeks, with an average gestational age of 28 weeks.

The ultrasound machine used in this study was a Voluson E8 or E10 (GE Healthcare Ultrasound) with a 4-8MHz transabdominal 2D transducer. In each examination, the biparietal diameter, head circumference, abdominal circumference, femur and other parameters of the fetus were measured to comprehensively evaluate the biological gestational age of the fetus, and then the screening of the structural malformation of each fetal system was conducted.

When detailed screening of fetal central nervous system structures, the transverse diameter of cerebellum and frontal lobe were measured by trans-cerebellar section. Standard trans-cerebellar section^4^: the probe clearly showed the midline of the brain, cavity of septum pellucidum, thalamus, bilateral cerebellar hemispheres and the vermis of the cerebellum, posterior fossa and Sylvian fissure. The measurement method was shown in Figure 1. The transverse diameter of cerebellum refers to the distance between the bilateral cerebellar hemispheres. The transverse diameter of frontal lobe refers to the distance between the medial margins of the bilateral skulls on the vertical line perpendicular to the bilateral skulls through the junction of the corpus callosum and the anterior margin of the septum pellucidum. The ratio of the transverse diameter of cerebellum to the transverse diameter of frontal lobe was automatically calculated using the software calculation package on the instrument. Each patient was measured by one experienced sonographer, and each was measured three times. When the fetus had poor image acquisition due to positional limitations, the pregnant woman was instructed to perform the scan after moderate activity. SPSS 26.0 statistical software was used to analyze and process the data.

**Fig. 1.**
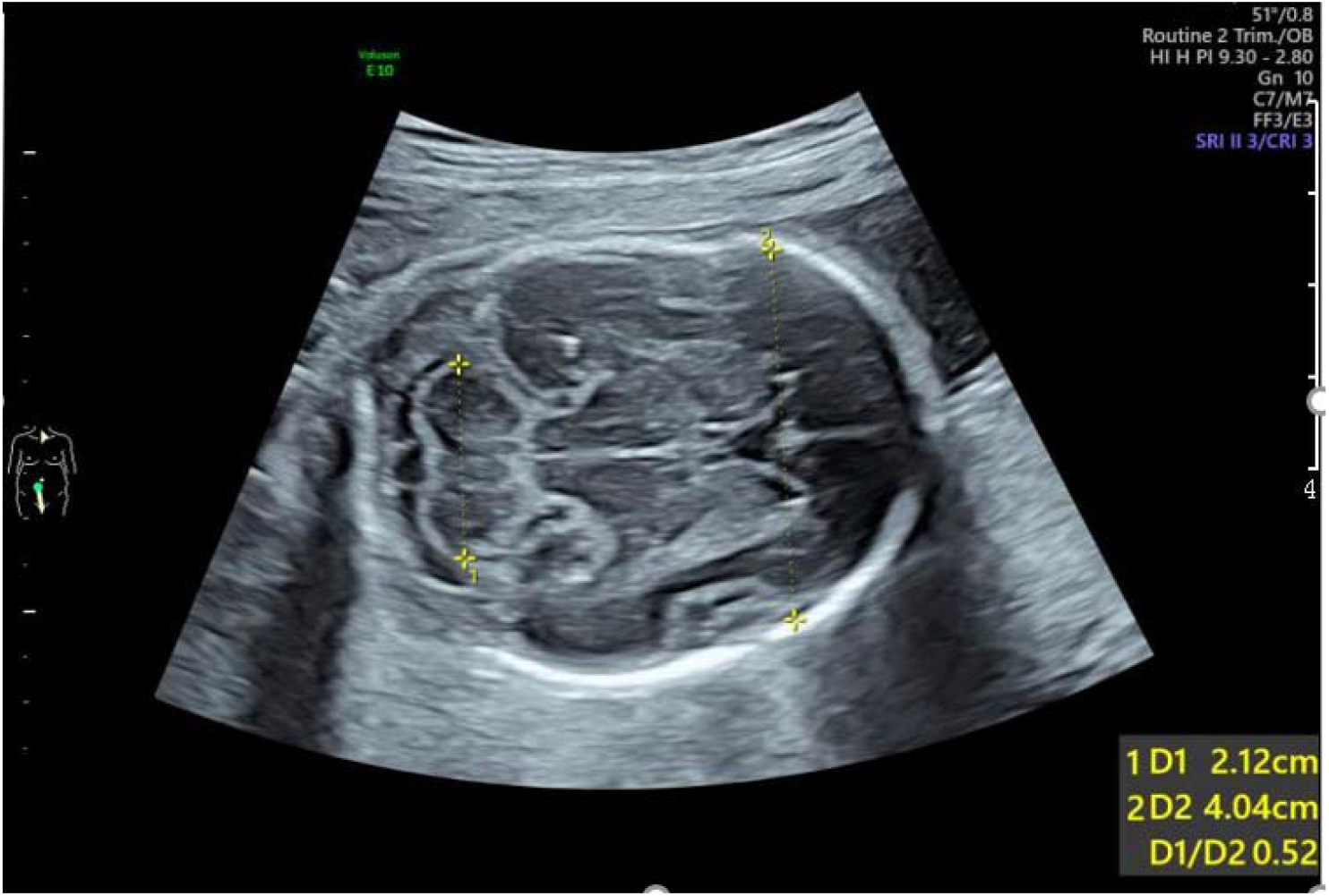
Measurement of the transverse diameter of cerebellum and transverse diameter of frontal lobe at 24 weeks of gestation. 1: transverse diameter of cerebellum; 2: transverse diameter of frontal lobe

The main steps were as follows:(i) All measured data were counted and divided into 16 groups according to gestational age. The gestational age sample size of each group was counted, the mean value and standard deviation of each parameter in each group 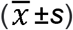 were calculated;(ii)the normality of each parameter value was tested by the Anderson-Darling test; (iii) for data that conformed to a normal distribution, the 95% reference range was calculated using the two-sided boundary value normal distribution method; for data with a skewed distribution, the normal reference range was calculated using the two-sided boundary value percentile method. The scatter plot of the relationship between the ratio of the transverse diameter of cerebellum to the transverse diameter of frontal lobe and the week of gestation was drawn, and a linear regression equation was established by linear regression analysis, with *p*<0.05 being statistically significant.

## RESULTS

A total of 870 subjects were included in the study. The sample data of the ratio of the transverse diameter of cerebellum to the transverse diameter of frontal lobe at different gestational weeks were consistent with or approximately normal distribution. The mean value, standard deviation and 95% reference range of each parameter were calculated at different gestational weeks, and the bilateral boundary value was taken. The data were shown in Table 1.

**Table 1.** The mean value, standard deviation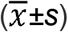 and 95% reference range of the ratio of the transverse diameter of cerebellum to frontal lobe at different gestational weeks

| Gestational weeks | Numbers | ratio of the transverse diameter of cerebellum to frontal lobe |  |
| --- | --- | --- | --- |
| | | ($\bar{x}\pm s$) | 95% reference range |
| 21 | 61 | 0.52±0.03 | 0.46~0.59 |
| 22 | 79 | 0.53±0.03 | 0.47~0.57 |
| 23 | 98 | 0.53±0.03 | 0.49~0.59 |
| 24 | 96 | 0.53±0.03 | 0.48~0.58 |
| 25 | 49 | 0.53±0.04 | 0.48~0.61 |
| 26 | 45 | 0.52±0.03 | 0.48~0.59 |
| 27 | 41 | 0.54±0.03 | 0.49~0.61 |
| 28 | 65 | 0.54±0.03 | 0.47~0.61 |
| 29 | 45 | 0.55±0.04 | 0.50~0.61 |
| 30 | 53 | 0.55±0.03 | 0.50~0.62 |
| 31 | 44 | 0.54±0.04 | 0.45~0.60 |
| 32 | 47 | 0.56±0.04 | 0.46~0.64 |
| 33 | 43 | 0.57±0.04 | 0.50~0.66 |
| 34 | 37 | 0.58±0.05 | 0.52~0.67 |
| 35 | 35 | 0.57±0.03 | 0.50~0.61 |
| 36 | 32 | 0.57±0.04 | 0.50~0.64 |

A scatterplot of the relationship between the ratio of the transverse diameter of cerebellum to the transverse diameter of frontal lobe and gestational weeks was drawn (Figure 2). Correlation analysis showed that the ratio of the transverse diameter of cerebellum to the transverse diameter of frontal lobe was positively correlated with gestational age, with a correlation coefficient of 0.413 (P < 0.001).

**Fig. 2.**
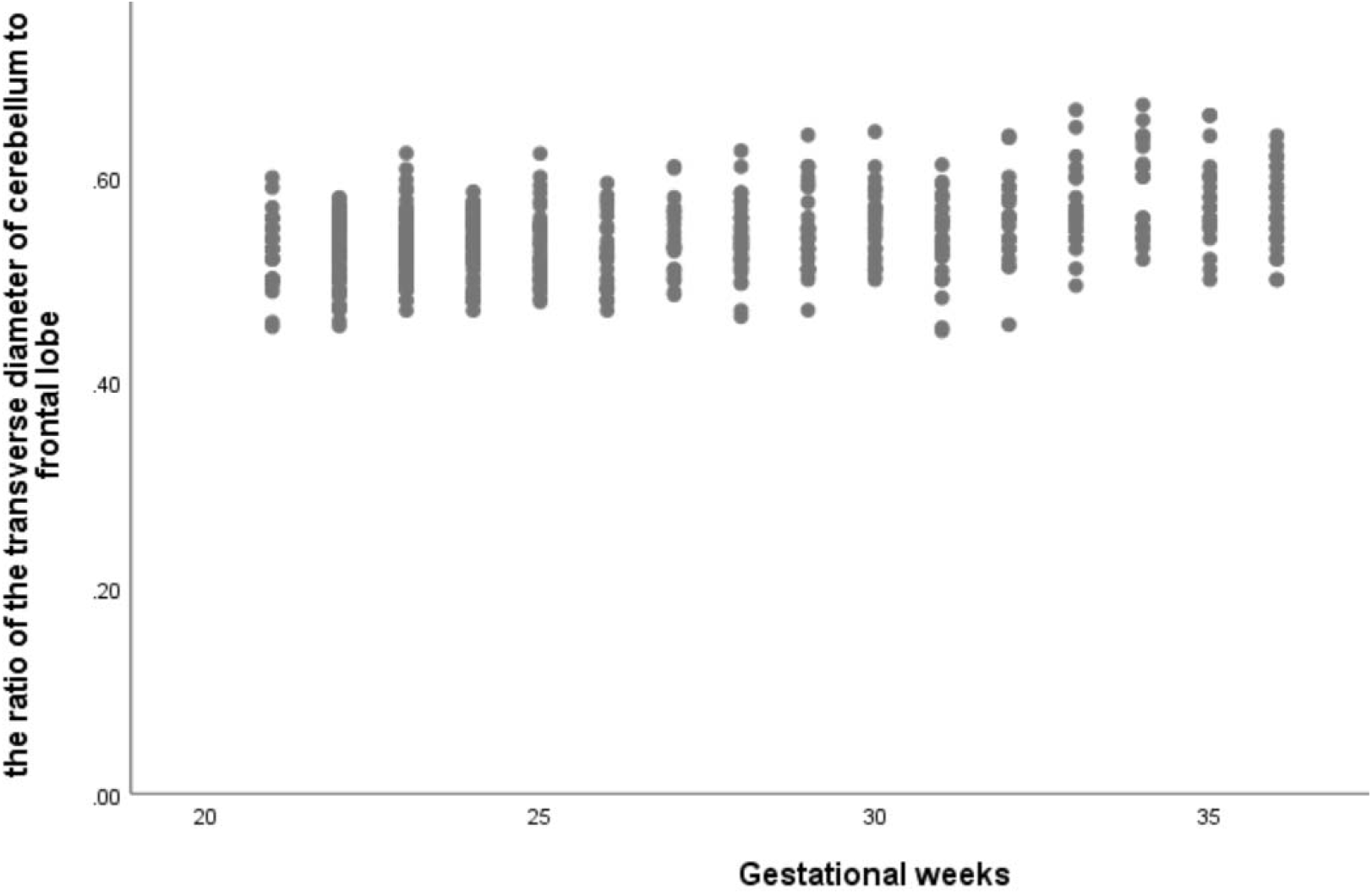
Scatterplot of the relationship between the ratio of the transverse diameter of cerebellum to frontal lobe and gestational weeks

Taking gestational weeks as the independent variable, the ratio of the transverse diameter of cerebellum to the transverse diameter of frontal lobe was used as the dependent variables for linear regression analysis. The result showed that there was a linear regression relationship between gestational weeks and the ratio of the transverse diameter of cerebellum to the transverse diameter of frontal lobe. The obtained linear regression equations was the ratio of the transverse diameter of cerebellum to the transverse diameter of frontal lobe =0.004× (gestational weeks-21)+0.530.

## DISCUSSION

Microcephaly is mainly manifested by delayed development of the frontal lobe. At present, there have been reports on the development rules of parameters related to fetal frontal lobe development^5-8^, which is helpful for the diagnosis of prenatal microcephaly. Since the frontal lobe is more involved in microcephaly than the cerebellum, the ratio of cerebellum to frontal lobe related measurement parameters should theoretically increase in the presence of microcephaly. Persutte^9^ established the reference value range of the ratio of transverse cerebellar diameter to anteroposterior frontal diameter in 221 normal fetuses, and found that the ratio of transverse cerebellar diameter to anteroposterior frontal diameter in microcephaly and trisomy 21 fetuses was above the 95th percentile, indicating that the developmental retardation of frontal lobe existed in microcephaly and trisomy 21 fetuses. Our previous study also confirmed that there was a certain regularity between the normal development of the frontal lobe and gestational week, and found that the transverse diameter of the frontal lobe had a higher correlation with gestational week than the anteroposterior diameter of the frontal lobe. Therefore, in the context of previous studies, our research team planned to further explore the relationship between the ratio of the transverse diameter of cerebellum and the transverse diameter of frontal lobes in normal mid-late pregnancy, and establish the normal reference range of the ratio of the transverse diameter of cerebellum to the transverse diameter of frontal lobe in different gestational weeks, so as to provide valuable information for the prenatal diagnosis of fetal microcephaly.

In this study, trans-cerebellar section was selected as the standard measurement section based on the following points. Firstly, trans-cerebellar section is one of the conventional sections for screening fetal central nervous system malformations; secondly, some studies have reported the relationship between the ratio of transverse cerebellar diameter to anteroposterior frontal diameter and gestational week. The anteroposterior frontal diameter was measured on the trans-thalamic section, so the ratio should be measured both in the trans-thalamic section and trans-cerebellar section, which increased the time of examination and measurement; thirdly, our previous study found that trans-cerebellar section showed the Sylvian fissure more clearly than trans-thalamic section, and Sylvian fissure is one of the important indicators for evaluating brain development as well. The result of this study showed that the ratio of transverse cerebellar diameter to transverse frontal diameter increased with gestational weeks in normal mid-late pregnancies. Correlation analysis showed that the correlation coefficient between the ratio of transverse cerebellar diameter to transverse frontal diameter and gestational age was 0.413(p<0.005). The result showed that there was a linear regression relationship between gestational weeks and the ratio of the transverse diameter of cerebellum to the transverse diameter of frontal lobe. The obtained linear regression equations was the ratio of the transverse diameter of cerebellum to the transverse diameter of frontal lobe =0.004× (gestational weeks-21)+0.530. The mean value, standard deviation and the 95th percentile of the ratio was calculated. If the ratio of the transverse diameter of cerebellum to the transverse diameter of frontal lobe is above the 95th percentile, the possibility of microcephaly should be highly alerted (Figure 3). The MRI examination of the fetus confirmed cortical dysplasia.

**Fig. 3.**
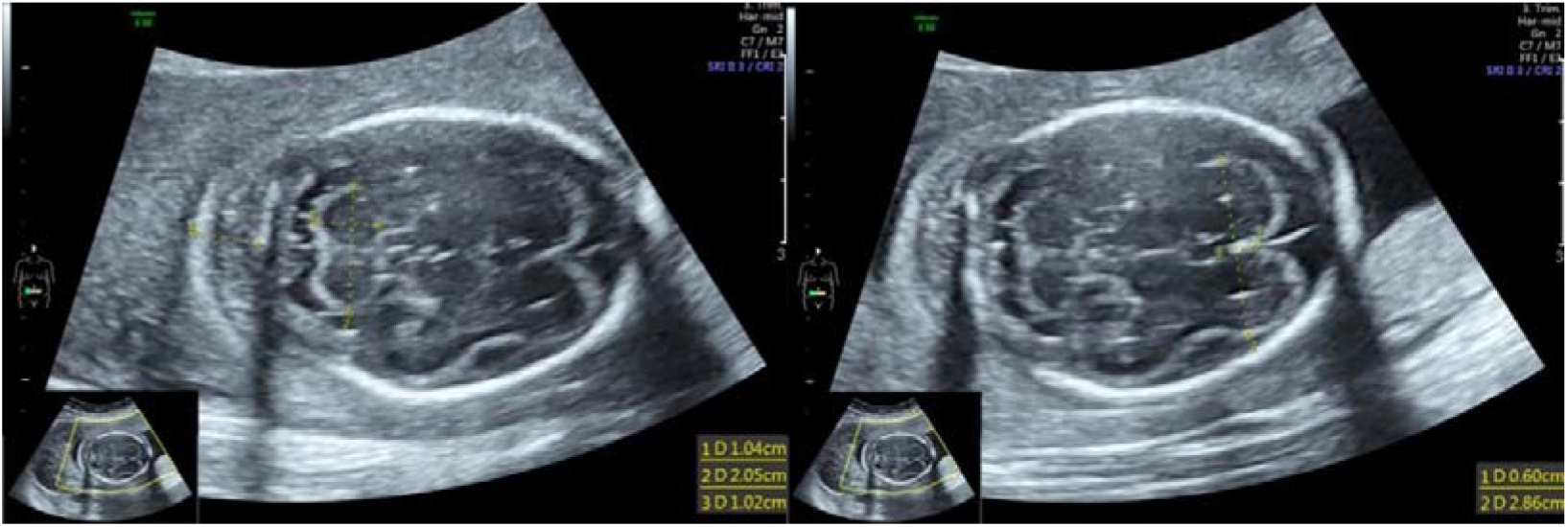
fetal head circumference < 3SD and the ratio of transverse cerebellar diameter to transverse frontal diameter was 0.72(> 95th percentile) at23 weeks of gestation

This study preliminarily established the normal reference value range of the ratio of the transverse diameter of cerebellum to the transverse diameter of frontal lobe in normal mid-late pregnancy fetus. If the ratio is above the 95th percentile, high vigilance should be given to the possibility of microcephaly.

## Data Availability

Data availability. The full data that supports the findings of this study are available from the corresponding author upon reasonable request.

## ACKNOWLEDGEMENTS

The authors thank all the women who participated in the study and acknowledge their significant contribution. Thank you to Zhang Dirong and Shi Yu for helping to design the research methods, Kong Fengbei and Yao Chunxiao for support with statistical analysis, She Ying and Wu Guoru for proposing some strategies for writing.

